# Antibiotic stewardship in premature infants: a systematic review

**DOI:** 10.1101/2020.04.10.20060988

**Authors:** Polona Rajar, Ola D. Saugstad, Dag Berild, Anirban Dutta, Gorm Greisen, Ulrik Lausten-Thomsen, Sharmila S. Mande, Sushma Nangia, Fernanda C. Petersen, Ulf R. Dahle, Kirsti Haaland

**Affiliations:** Department of Pediatrics, Oslo University Hospital, Ullevaal, Oslo, Norway; Institute of Oral Biology, University of Oslo, Oslo, Norway; Department of Pediatric Research, University of Oslo, Oslo, Norway; Department of Infectious Diseases, Oslo University Hospital, Oslo, Norway; Institute of Clincal Medicine, Faculty of Medicine, Oslo University; TCS Research, Tata Consultancy Services Ltd., Pune, Maharashtra, India; Copenhagen University Hospital Rigshospitalet, Copenhagen, Denmark; Lady Hardinge Medical College and Kalawati Saran Hospital, New Delhi, Delhi, India; Centre for Antimicrobial Resistance, Norwegian Institute of Public Health, Oslo, Norway

## Abstract

Antibiotic treatment in premature infants is often empirically prescribed, and practice varies widely among otherwise comparable neonatal intensive care units. Unnecessary and prolonged antibiotic treatment is documented in numerous studies. Recent research shows serious side effects and suggests long-term adverse health effects in prematurely born infants exposed to antibiotics in early life. One preventive measure to reduce unnecessary antibiotic exposure is implementation of antibiotic stewardship programs. We reviewed the literature on implemented antibiotic stewardship programs focusing on premature infants. Six academic databases were systematically searched and eleven articles met inclusion criteria. Articles were grouped according to common area of stewardship actions; 1) Focus on reducing initiation of antibiotic therapy, 2) Focus on shortening duration of antibiotic therapy, 3) Various infrastructural stewardship implementations. The studies differed in their cohort composition and measured outcomes. We provide an overview of the reduction in antibiotic use achieved. Antibiotic stewardship programs are effective especially when they use a multifactorial approach and are tailored to premature infants. Implementation of antibiotic stewardship programs targeting premature infants should be considered in all neonatal intensive care units. The Norwegian Research Council (project number 273833) and the Olav Thon Foundation supported the study.

## Introduction

Already in the first half of the twentieth century, in the wake of antibiotic discoveries, P. Ehrlich, A. Fleming and others warned about the possibility that microbes could develop resistance to antibiotics.^1^ A number of new antibiotics were developed through the 1950s and 1960s, but few novel antibiotics have been introduced to the market since.^2^ Unfortunately, the battles against drug resistant organisms are becoming increasingly challenging.^3,4^ Simultaneously, unparalleled progress in health care services has augmented the expectations to treatment. Immunosuppressive drug treatments and invasive surgical procedures to improve life quality of an aging population are now common practice, and rely on the existence of effective antibiotics. Similarly, the treatment and survival of newborn infants, in particular the premature, often rely on effective antibiotics. Sepsis and meningitis are leading causes of death and morbidity, contributing to 15 % of neonatal deaths worldwide (2017).^5^

Early onset sepsis (EOS) is defined by positive microbial cultures from blood or cerebrospinal fluid (CSF) obtained within 72 hours after birth, and late onset sepsis (LOS) after 72 hours.^6^ However, blood cultures are often falsely negative due to difficulties in obtaining sufficient blood sample, low bacteremia levels and use of intrapartum antibiotic prophylaxis. A functional diagnosis of sepsis is thus more frequently applied, by taking into account risk factors, clinical status and other laboratory parameters of the infant.^7^ As laboratory tests alone are not sufficient and clinical signs can be prone to subjective interpretation, risk assessment with a low threshold is often used for starting empiric antibiotic therapy.^7,8^ Also, non-blood stream infection is poorly defined and diagnosed in preterm infants in current neonatal practice. Thus, while blood culture results are important in guiding the decision to continue, change or discontinue antibiotic treatment, they are of less value to guide the decision of when to start antibiotic therapy.

Incidence of EOS is inversely proportional to gestational age (GA) at birth.^8^ Mortality rates from EOS are higher at lower GA and in low birth weight infants (Figure 1).^8^ Unreliable clinical signs, disastrous outcome in case of delayed start of antibiotic treatment, and reluctance to withdraw initiated treatment often result in overuse of antibiotics in neonatal intensive care unit (NICU).

**Figure 1:**
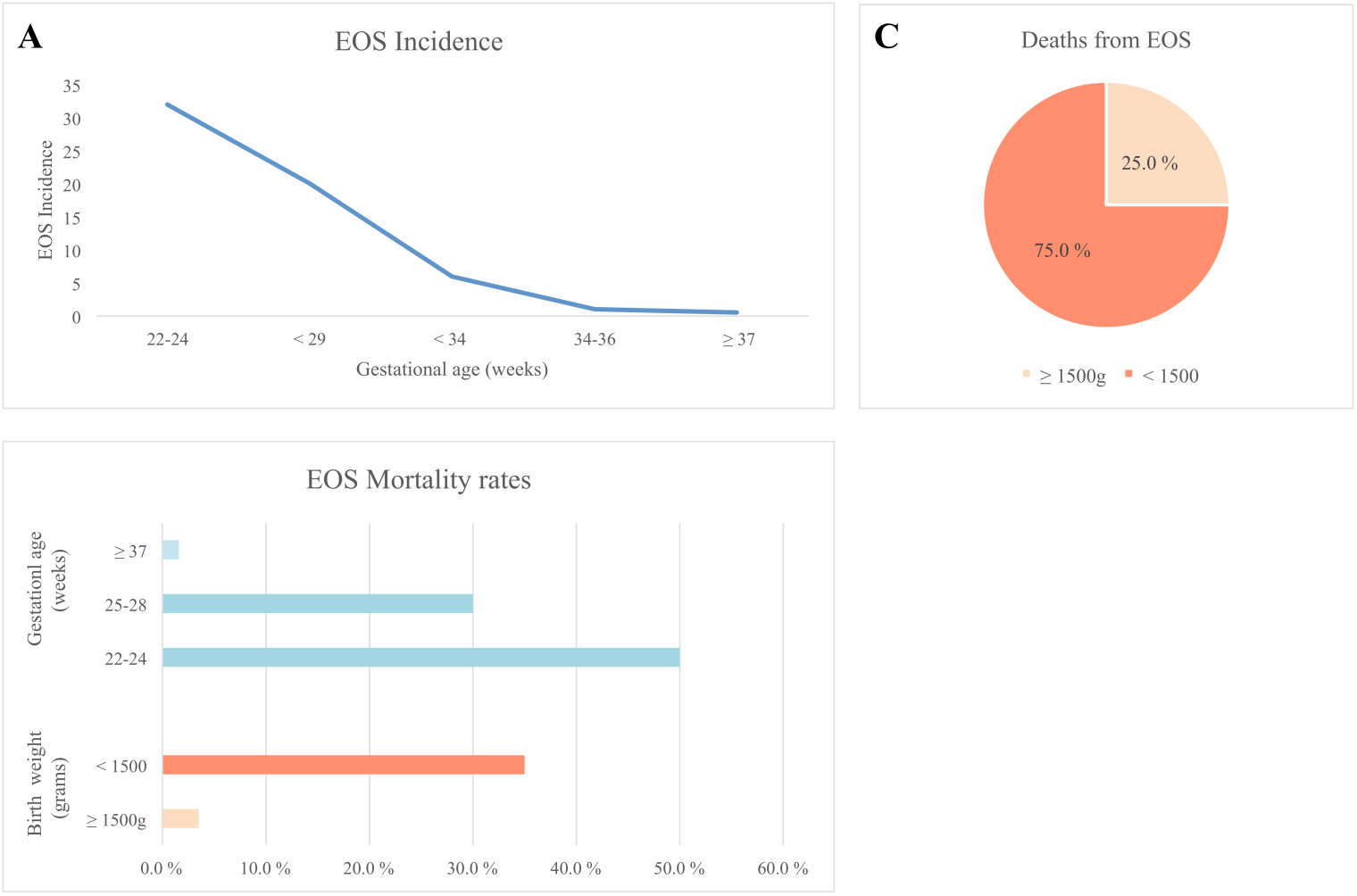
The incidence and mortality rates for early onset sepsis (EOS). (A) Incidence of early onset sepsis (EOS) at gestational age (GA)^9-13^: 0.5/1000 at ≥ 37 weeks, 1/1000 at 34 - 36 weeks, 6/1000 at < 34 weeks, 20/1000 at < 29 weeks, 32/1000 at 22 - 24 weeks. (B) Mortality rates from EOS at GA^10-12^: 1.6% at ≥ 37 weeks, 30 % at 25 – 28 weeks, 50 % at 22 – 24 weeks and at birth weight^9^: 3.5 % at ≥ 1500 g, 35 % at < 1500. (C) Death from EOS at birth weight: 75 % of deaths from EOS occur amongst very low birth weight (VLBW < 1500 g) infants.^9^

## Background

The term “antibiotic stewardship program” (ASP) was first introduced in 1996 and has since shown an exponential increase in popularity.^14^ MacDougall et al. defined ASP as “on-going efforts by a health care organization to optimize antimicrobial use among hospitalized patients in order to improve patient outcomes, ensure cost-effective therapy, and reduce adverse sequelae of antimicrobial use (including antimicrobial resistance (AMR))”.^15^

Antibiotics, the most prescribed medicines in NICU settings, are essential to a large proportion of premature infants.^16,17^ A risk-based approach with low threshold is often used for starting antibiotic treatment right after birth, an approach that has successfully lowered EOS incidence, but increased number of non infected infants exposed to antibiotics.^6^ Empiric therapy is often extended to five to seven days even in the absence of positive blood cultures.^18^

Antibiotic treatment for more than five days in infants with negative blood cultures is associated with increased risk of necrotizing enterocolitis (NEC), bronchopulmonary dysplasia (BPD), invasive fungal infections (IFI), retinopathy, periventricular white matter damage and death.^19,20^ Each additional day of antibiotic exposure in the absence of positive blood cultures elevates the risk of NEC in very low birth weight (VLBW) babies 7 – 20 %.^21,22^ Birth mode, feeding, skin contact and long stays in a hospital environment are some of the factors modulating the development of infant’s microbiome.^23^ The infant’s developing microbiome increases in richness and diversity from the beginning of life and is particularly susceptible for modulating factors in infancy.^24,25^ By the age of two or three years it resembles the adult microbiome.^26^ Perturbations occurring early in life influence host physiology, biological mechanisms involved in maturation of immune system and neurodevelopment.^27^ In addition, antibiotic disruption of the microbiome may carry lasting consequences reflected as dysbiosis, increased carriage of antibiotic resistance genes (ARG) and multidrug resistant organisms.^28,29^ Active efforts to reduce immediate and long-term adverse effects should be strived for. Decreasing prolonged antibiotic exposure could be a significant factor for improving infant’s long-term health outcomes.

### Current state

In a recent study, Flannery et al. (2018)^30^ demonstrated that the majority of very-low-birth-weight (VLBW, < 1500 g) and extremely-low-birth-weight (ELBW, < 1000 g) infants from nearly 300 USA hospitals were treated with antibiotics in their first days of life (78·6 % and 87·0 %, respectively). Additionally, 26·5 % VLBW and 37·8 % ELBW infants received more than five days of antibiotic treatment. Importantly, there were major differences in duration of antibiotic therapy between hospitals that could not be explained solely by medical reasons. Also in Norway, > 50 % of infants born at GA < 32 weeks received intravenous antibiotics within the first 14 days of life in the period from 2015 to 2018.^31^ In the period from 2009 to 2011 median treatment duration (interquartile range) was 8 (7-10) and 6 (5-7) days for culture positive and culture negative EOS, respectively.^31^

Antibiotic use is imperative and should be expected to remain high in premature infants, but unnecessary antibiotic exposure should be minimized due to substantial risks of adverse effects.^32^ This review aims to summarize available knowledge on implemented ASPs in neonatal intensive care units, focusing on premature infants.

## Methods

This systematic review was performed using all applicable items from the PRISMA guidelines for reporting of systematic reviews.^33^

We performed a search in the following databases: PubMed (Medline), McMaster PLUS, Cochrane Database of Systematic reviews, UpToDate, Cochrane central register of Controlled trials and National Institute for Health and Care Excellence, including all articles published to 05.12.2019. Full search terms and search strategy are provided (Appendix 1). Due to the novelty of APS in settings with premature infants, the term “neonatal infant” was included in the search process. No previous systematic review of ASPs in premature infants was identified.

We retrieved 1210 tittles, no duplicates were found. Three authors (PR, ODS, URD) screened the titles and abstracts of all (1210) studies identified through the search and excluded comments and guidelines. Papers that did not include premature patients were also excluded, as were papers where antibiotic stewardship actions were directed towards microorganisms (reducing infections with multiresistant organisms, monitoring ARG) and papers that only reported the current state of antibiotic consumption and possibilities for ASP, but lacked the implementation and evaluation of ASP actions (Figure 2). In total, 27 full-text articles were retrieved for all studies that met the inclusion criteria, or required more information than was provided in the abstract for an informed decision. Two investigators (PR, KH) independently assessed the full-text articles. A total of 11 papers were included in the review (Table 1), reasons for excluding remaining 16 studies are described (Appendix 2).

**Table 1:** Included studies. *Grouping according to type of ASP actions used in the studies, explained in results. † SMAP = Scores for neonatal acute physiology with perinatal extension - II assessment. ASP = antibiotic stewardship program. CDC = Centers for Disease Control and Prevention. DOT = days of therapy. DOL = days of life. DDD = defined daily dose. ELBW = extremely low birth weight. EOS = early onset sepsis. GA = gestational age. LBW = low birth weight. LOS = late onset sepsis. MALDI- TOF = Matrix-Assisted Laser Desorption/Ionization-Time Of Flight. No. = Number of. VLBW = very low birth weight.

| Source | Location | No. infants | Gestational age of participates | Study details, interventions | Measure of outcome | Relevant findings | Group* |
| --- | --- | --- | --- | --- | --- | --- | --- |
| 1. Achten et al. (2018)<sup>34</sup> | The Netherlands | 208 | ≥ 35 weeks GA, with either elevated maternal EOS risk or/and possible EOS based on clinic presentation within 72 hours. | Implementation of sepsis calculator to reduce empiric antibiotics for suspected EOS. | Reduction in empirical use of antibiotics. | Reduced empiric antibiotic therapy for suspected EOS after sepsis calculator implementation (from 4·8 % to 2·7 %, $p < 0·001$). | 1 |
| 2. Astorga et al. (2018)<sup>16</sup> | Wisconsin, USA | 1,203<br>(564 pre- ASP) | 143 (25·4 %) pre- and 152 (23·8 %) post-ASP infants with GA < 34 weeks. 133 VLBW infants, 11 % in the pre- and 11·1 % in the post-ASP group. | Retrospective before-and-after double-cohort observational study, evaluating implementation of an automatic 48-hour electronic stop on all parenteral antibiotics. | DOT/1000 patient-days. | No difference in initiation of antibiotics. Total doses of antibiotics per patient decreased by 35 % ($p < 0·0001$). Total antibiotic doses per patient-day decreased by 25 % ($p < 0·0001$). | 2 |
| 3. Jinka et al. (2017)<sup>35</sup> | Andhra Pradesh, India | 2,452 (1176 pre-ASP) | No GA data.<br>LBW infants: 406 (35 %) pre-, 480 (37 %) post-ASP.<br>VLBW infants: 106 (9 %) in pre-, 74 (6 %) post-ASP. | A new protocol for empiric therapy of neonatal sepsis based on review of blood culture susceptibility data.<br>Limitations: No individual patient data. | DDD/100 patient-days. | The proportion of infants on antibiotics decreased ($p < 0·001$), but there was no significant change in consumption of antibiotics after ASP implementation when adjusted by average weight per month. | 3 |
| 4. Kitano et al. (2019)<sup>36</sup> | Nara, Japan | 1,107 (913 pre-ASP) | Average GA of 36 weeks. 10·4 % and 10·3 % VLBW; 1·5 % and 2 % ELBW (pre- and post-ASP, respectively) | Implemented a protocol of antimicrobial treatment at NICU: new start and stop criteria for antibiotic therapy, weekend report of blood culture results, stopping ordering antimicrobials for the next day. | DOT/1000 patient-days. | After ASP implementation, DOT/1000 patient-days decreased for 76·2 % ($p < 0·001$), as did the percentage of neonates receiving any antibiotic therapy and the percentage of neonates receiving prolonged therapy ($p < 0·001$ for both). | 1/2/3 |
| 5. Lu et al. (2019)<sup>37</sup> | Shanghai, China | 13,540<br>(7,754 pre- ASP) | 4,497 (58 %) infants baseline and 3,502 (60 %) post-ASP had GA < 37 weeks. | SMAP† implementation, baseline and post-intervention assessments. SMAP consisted of antibiotic restrictions, reviews of electronic medical records, SNAPPE-II assessments and a 48-hour automatic stop order on empiric antibiotic therapy. | DOT/1000 Patient days. | 33 % decrease in DOT/1000 patient days ( $p = 0·0001$ ).<br>A significant decrease in LOS ( $p = 0·03$ ), in percentage of infants with discontinued treatment after 48 hours ( $p = 0·0001$ ) and in percentage of infants with neg. blood cultures treated for ≥ five days ( $p = 0.02$ ). | 2/3 |
| 6. McCarthy et al. (2018)<sup>38</sup> | Cork, Ireland | 312 (124 baseline, 82 1<sup>st</sup> intervention, 106 2<sup>nd</sup> intervention) | Infants with GA < 37 weeks: 43 (35 %) baseline, 39 (47.5 %) 1<sup>st</sup> intervention, 26 (25 %) 2<sup>nd</sup> intervention | A prospective audit was first preformed, followed by implementation of electronic prescribing and staff education. Two subsequent audits were preformed, with a 36-hour automatic stop of antibiotic therapy implemented after the second re-audit. | DOT/1000 patient-days and infants receiving prolonged antibiotic therapy. | 27 % decrease in DOT/1000 patient-days, as well as significant decreases of prolonged (> 36 hours) antibiotic therapy in negative sepsis evaluations ($p = 0.0040$) and decreased prolonged (> 5 days) antibiotic treatment in culture negative sepsis ($p = 0.0009$). | 2/3 |
| 7. Nitsch-Osuch et al. (2015)<sup>39</sup> | Wroclaw, Poland | 418 (208 pre-ASP) | No GA data. | Implementation of hospital antibiotic policy by the hospital infection control team (reassigning 1<sup>st</sup>, 2<sup>nd</sup> and 3<sup>rd</sup> line antibiotics).<br>Limitations: No individual patient data. | DDD/100 patient-days. | Slight increase in total antibiotic consumption, but an improved profile of antibiotic consumption. | 3 |
| 8. Nzegwu et al. (2017)<sup>40</sup> | Massachusetts, USA | 4,551 (1,204 pre-ASP) | Average GA was 35.4 weeks pre- and 35.5 weeks post-ASP.<br>ELBW: 118 (9.8 %) pre-, 282 (8.4 %) post-ASP. | New recommendations for evaluation and treatment of neonatal sepsis. Prescriber audit and feedback. | DOT/1000 Patient day. | Significant decline in LOS evaluation and prescription events post-ASP ($p < 0.001$). However, this was not reflected in the total antibiotic utilization, as only a slight decrease in DOT/1000 patient-days ($p = 0.699$) was observed. | 3 |
| 9. Tagare et al. (2010)<sup>41</sup> | Pune, India | 140 (71 control group) | All participants had GA ≤ 36 weeks.<br>VLBW: 53 (26 in control group). | A randomized controlled trial of routine antibiotic treatment.<br>Premature infants (with no other risk factors) were assigned to either control (no antibiotic therapy) or intervention group (five days of intravenous antibiotic treatment). | Incidence of sepsis. | Comparable incidence of sepsis in both groups (25.4 % and 31.9 % for control and intervention group, respectively), also in VLBW infants (42.3 % and 59.3 % for control and intervention group, respectively). | 1 |
| 10. Ting et al. (2019)<sup>42</sup> | Vancouver, Canada | 2,670 (2003 pre-ASP) | VLBW: 569 pre-ASP, 154 post-ASP. | A retrospective audit focused on prolonged (> three days) antibiotic prescriptions and evaluation according to the 12 steps of CDC. Audit as repeated 12 months after ASP implementation. New approach to LOS management in order to reduce unnecessary prolonged antibiotic exposure.<br>New diagnostic tools (MALDI-TOF) for early identification of organisms, education of staff. There was no change in the protocols for empiric antibiotic therapy (except restriction of linezolid). | Inappropriate antibiotic-days/1000 patient-days. | Inappropriate antibiotic-days/1000 patient-days decreased from 3.56 to 1.73 [RR, 0.49 (95% CI: 0.33–0.71)], but not improvement was seen in the VLBW group. | 2/3 |
| 11. Tolia et al. (2017)<sup>43</sup> | Texas, USA | 674 (313 pre-ASP) | 101 infants (32·3 %) pre- and 114 infants (31·6 %) post- ASP with GA ≤ 26 weeks.<br>All were VLBW. | Automatic 48-hour electronic stop and education of health staff.<br>Limitation: antibiotic use was recorder only for the first 7 DOL. | DOT/1000 Patient days and percentage of infants receiving prolonged therapy. | 28 % decrease in DOT/1000 patient days (p < 0·001)<br>Decrease in percentage of infants with antibiotics > 48h (p < 0·001). | 2 |

**Figure 2:**
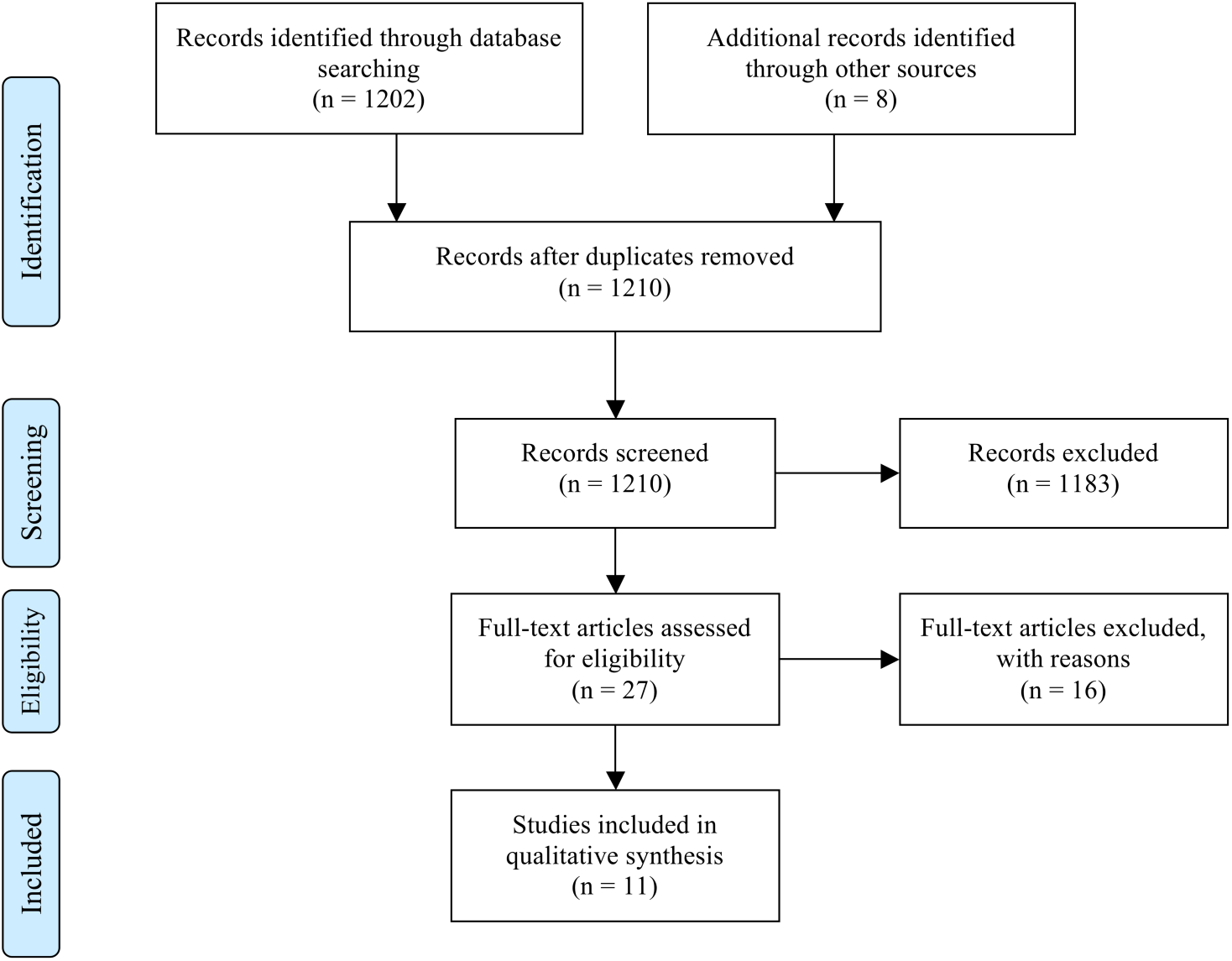
Study selection process.

## Results

The 11 selected articles varied greatly in their study population, interventions and measured outcomes. To compare their findings we identified I - common actions (Figure 3) and II - common units of measure for reporting results (Figure 4 - 5).

**Figure 3:**
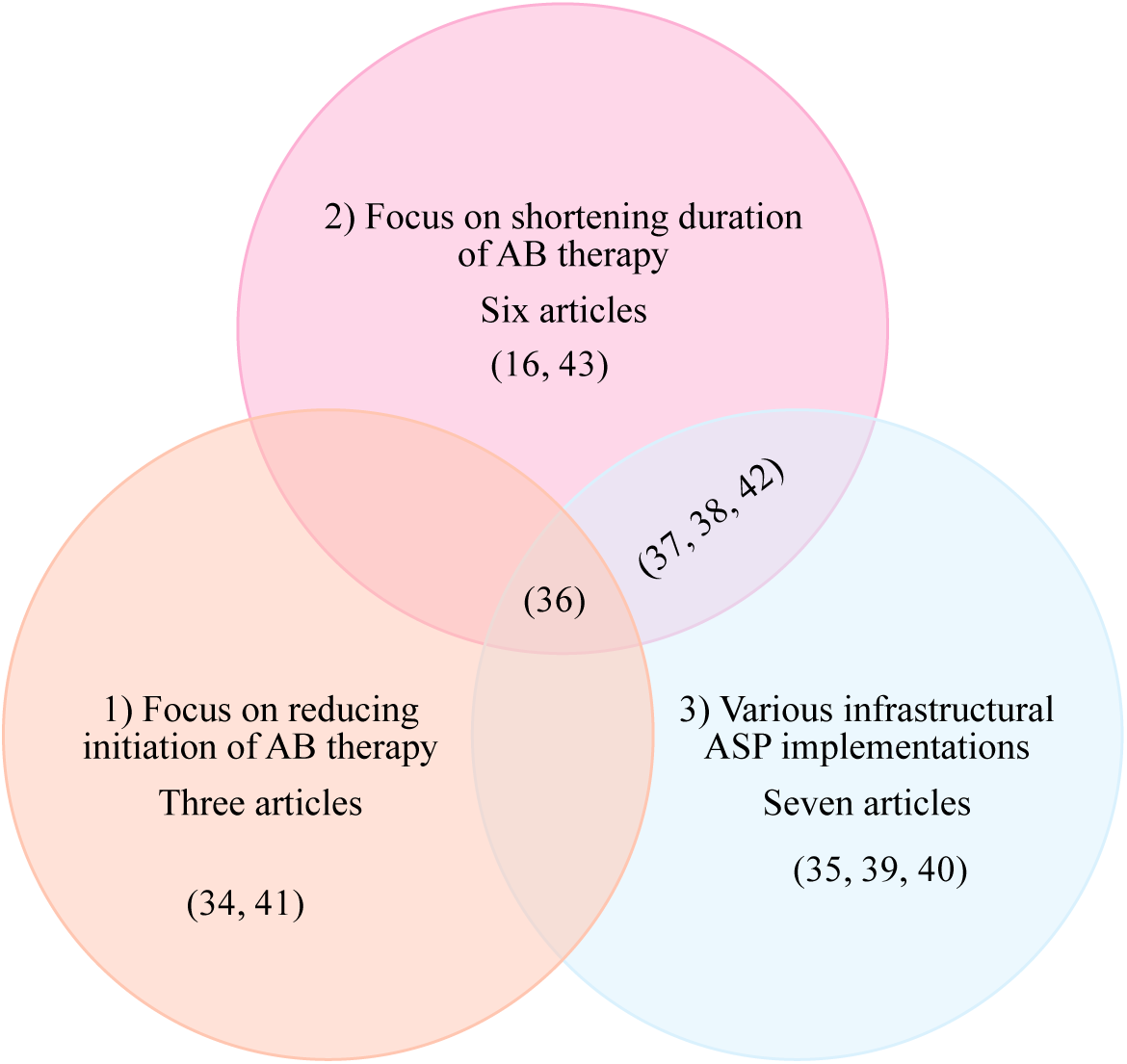
I Antibiotic stewardship interventions grouped according to common areas of actions. 1) Focus on reducing initiation of antibiotic therapy, 2) Focus on shortening duration of antibiotic therapy, 3) Various infrastructural ASP implementations. There is overlap in the studies implementing various actions. The first two areas used actions tailored to NICU patients, while actions from the third area are less specific and could be adapted to health settings in general.

**Figure 4:**
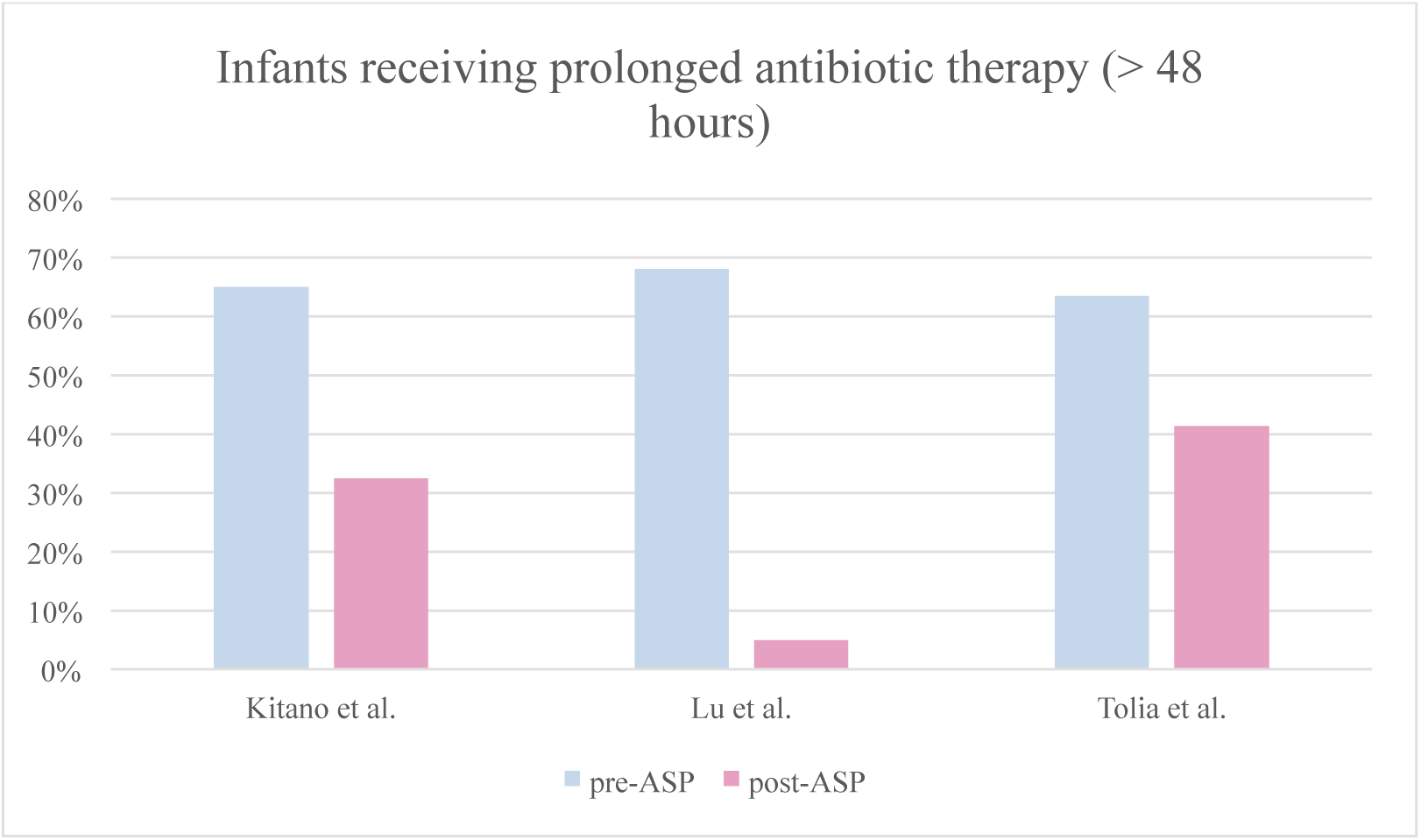
Percentage of infants receiving prolonged (> 48 hours) antibiotic therapy. Kitano et al. achieved a reduction of prolonged antimicrobial treatments from 65 % to 32·5 % (p < 0.001).^36^ Lu et al. had an increase in percentage of discontinued antibiotic courses ≤ 48 hours from 32 % to 95 % (baseline cohort of 7,754 infants and intervention cohort of 5,786 infants).^37^ There was also a decrease in infants with culture negative sepsis receiving ≥ 5 days of antibiotics (from 66 % baseline to 33 % post-intervention). Tolia et al. also achieved a lower percentage of infants with > 48 hours of antibiotic exposure from 63·4 to 41·3 % (p < 0·001), in a cohort of 313 and 361 infants, respectively.^43^

**Figure 5:**
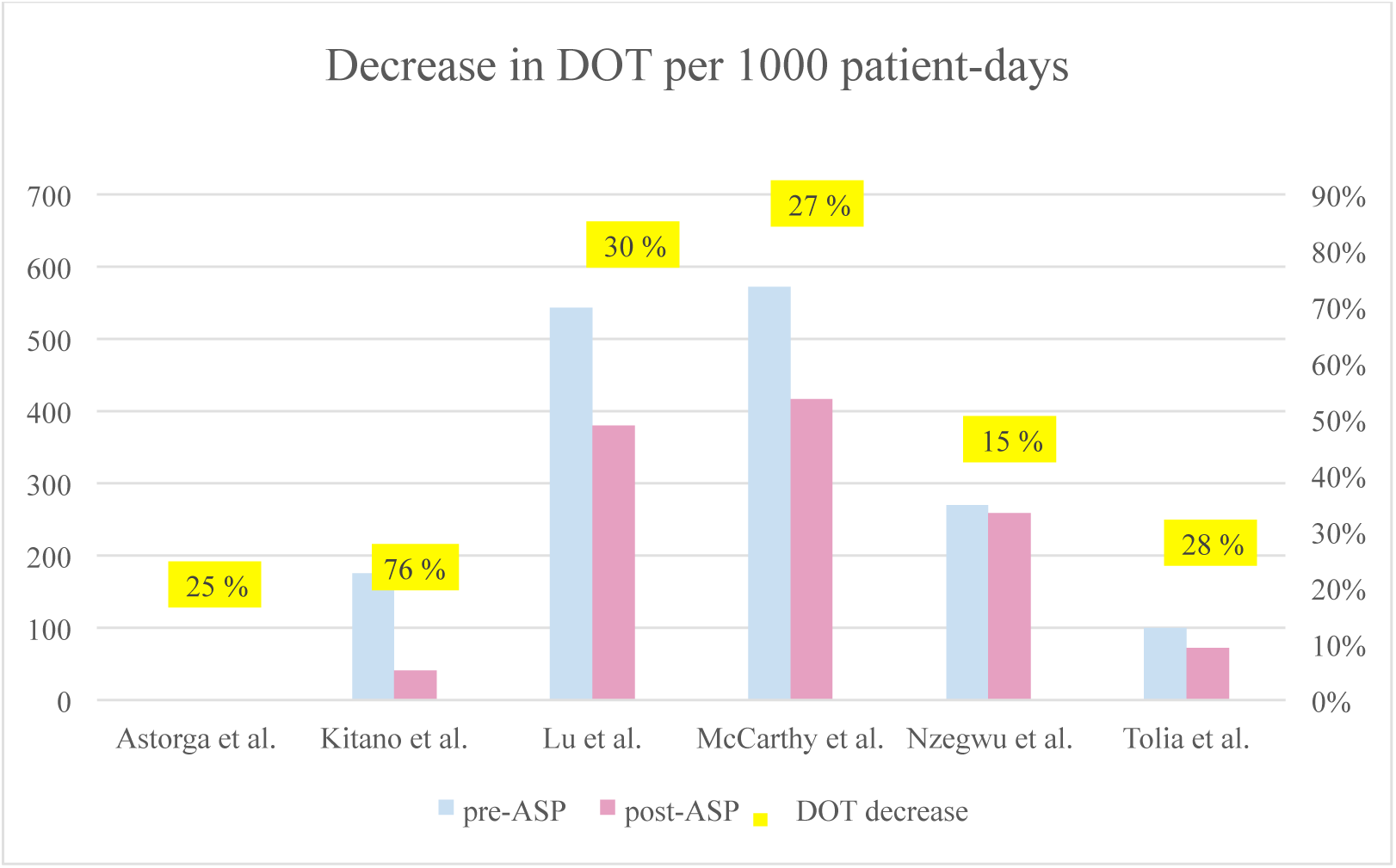
Days of therapy (DOT) /1000 patient-days. Astorga et al. was able to achieve a significant 25 % decrease (p < 0·0001) in a post intervention cohort of 639 infants (564 infants at baseline).^16^ Kitano et al. achieved a decrease from 175·1 to 41·6 DOT/1000 patient-days (76 %, p < 0·001).^36^ Lu et al. achieved a decrease from 543 to 380 DOT/1000 patient-days (30 %, p = 0·0001).^37^ McCarthy et al. reduced antibiotic use from 572 to 417 DOT/1000 patient-days (27 %, p = 0·0001) after 2^nd^ intervention.^38^ Nzegwu et al. did not find a significant decrease of total antibiotic consumption (p = 0·669), while consumption of ampicillin decreased significantly (p = 0·037) in a cohort of 4,551 infants (1,204 pre-ASP).^40^ Tolia et al. achieved a significant reduction from 99·5 DOT/1000 patient days (313 infants) to 71·7 DOT/1000 patient days (361 infants), a 28 % reduction in antibiotic use (p < 0·001).^43^

Most commonly used measures of units describing success of ASP were Days of therapy/1000 patient-days (DOT) and Defined daily dose (DDD).^44^ DOT represents the actual dose received by the patient, and is preferred in the paediatric patients as dosage is weight-adjusted. Papers listed under one of the first two action areas used individual patient data, and mostly expressed their results as DOT, or percentages of infants receiving prolonged or no antibiotic treatment after ASP implementation. Some papers in the third area reported their result as DDD which gives information of the total volume of antibiotic used by a department. DDD is a technical unit of measurement, convenient for comparing drug consumption between hospitals, or at an international level. DDD is normally assigned for 70 kg adults not optimized for use in children due to the variability of children’s doses. It is easy to obtain (pharmaceutical records), but lacks individual level data.

Group1) Studies focusing on reducing initiation of antibiotic treatment. The Kaiser sepsis calculator (http://kp.org/eoscalc^45^) to evaluate the risk for EOS in infants was used by Achten et al.^34^, while Tagare et al.^41^ performed a randomised clinical trial to evaluate the protective effect of empiric antibiotic coverage in premature infants in low risk situations. They randomised premature infants with no other risk for infections in two groups. The control group did not receive any antibiotic prophylaxis, while the intervention group received five days of prophylactic antibiotic treatment. Kitano et al. applied interventions from all three actions areas including new criteria for initiation of antibiotic treatment, based on maternal chorioamnionitis, infant’s clinical presentation and laboratory values.^36^

Group 2) To shorten duration of antibiotic treatment, Astorga et al. implemented a 48- hour automatic stop on empiric antibiotic therapy, initiated in infants at risk for infection according to the hospital protocol without other changes to their practise.^16^ Tolia et al. and Lu et al. implemented an electronic stop order at 48 hours in addition to other ASP actions.^37,43^ McCarthy et al. specifically targeted prolonged antibiotic courses in their second intervention period, implementing a hard stop for electronically prescribed antibiotics after 36 hours in asymptomatic infants with negative blood cultures and two consecutive CRP values within normal range.^38^ Similarly, Ting et al. encouraged discontinuation of antibiotic treatment within 48 hours in infants with negative blood cultures, no clinical suspicion for sepsis and serial CRP values in normal range.^42^ The ASP protocol of Kitano et al. included criteria for stopping antibiotics within 48 h in infants with negative blood cultures and without clinical suspicion of sepsis.^36^

Group 3) included various infrastructural ASP implementations. Jinka et al. and Nitsch-Osuch et al. implemented a protocol for empiric treatment and an ASP focusing mainly on restriction of antibiotic prescriptions, respectively.^35,39^ They provide no individual level data. Nzegwu et al. evaluated the implementation of new guidelines for neonatal infection assessment and unit-wide education on ASP, focusing especially on management of LOS, without any specific actions (such as an automatic stop order) taken.^40^ Nitsch-Osuch evaluated antibiotic consumption after new guidelines for prescribing antibiotics (including reassigning 1^st^, 2^nd^ and 3^rd^ line antibiotics).

Lu et al. reassigned 1^st^, 2^nd^ and 3^rd^ line antibiotic in order to restrict consumption of third generation cephalosporins, meropenem and linezolid.^37^ They reviewed electronic records of all antibiotic use in the NICU. Health personnel was informed and trained about the ASP interventions after the baseline period was concluded. McCarthy et al. focused on educational interventions based on monitored antibiotic prescribing data.^38^ Ting et al. focused on implementing three steps of the 12-steps program (CDC), adjusted for the NICU population; “target the pathogen”, “practice antimicrobial control”, “know when to say no”.^42^ After preforming a retrospective audit for episodes of prolonged (> three days) antibiotic prescriptions, their multidisciplinary ASP NICU team defined appropriate uses of antibiotics in different clinical situations, preformed staff education and implemented a new diagnostic tool to allow for earlier identification of organisms. Kitano et al. used action from all three areas.^36^ Their ASP protocol included new criteria for antibiotic initiation and discontinuation, specific to neonatal patients. Cases of non-compliance were discussed collegiately on daily basis. Other infrastructural ASP actions included availability of blood culture results also on weekends and holidays.

### II Antibiotic stewardship articles using common units of measure to report their results

#### Percentage of infants starting on antibiotics

Two studies (Achten et al., Kitano et al.) reported their results as the decrease of percentage of patients starting antibiotic therapy.^34,36^ Achten et al. achieved a reduction in percentage of infants treated with antibiotics from 4·8 % to 2·7 % of total births.^34^ Kitano et al. achieved a reduction from 55·3 % of infants pre-ASP to 20·6 % infants receiving any antibiotic treatment post-ASP implementation.^36^

#### Infants receiving prolonged (> 48 hours) antibiotic therapy

Three studies (Kitano et al., Lu et al., Tolia et al.) showed the efficiency of their ASP as decrease in percentage of infants receiving prolonged (> 48 hours) antibiotic therapy (Figure 4).^36,37,43^ McCarthy et al. showed significant reduction of prolonged (> 36 hours) therapy (p < 0·001).^38^ They achieved this after 2nd intervention, reducing prolonged courses from 82 DOT (baseline) and 79·8 DOT (after1^st^ intervention) to 7·5 DOT (124 infants in the baseline group, 82 and 106 infants in the 1^st^ and 2^nd^ intervention groups, respectively).

#### Days of therapy (DOT) /1000 patient-days

Six of the articles expressed their result in DOT/1000 patient-days (Figure 5).

#### Defined daily dose (DDD)/100 patient-days

Two papers, Jinka et al. and Nitsch-Osuch et al. used general oriented approaches for their ASP and expressed results as volumetric units for the whole NICU, lacking individual data.^35,39^ Jinka et al. observed a non-significant reduction of antibiotic consumption (from 14·47 to 11·47 DDD/100 patient-days, p = 0·57) in a cohort of 1,176 and 1,276 infants pre- and post-ASP, respectively.^35^ They did however achieve a significant increase in consumption of first line antibiotics (p < 0·001) and a significant decrease in 3^rd^ generation cephalosporins (p = 0·002). The effect of the ASP described by Nitsch-Osuch et al. resulted in an increase of antibiotic consumption (from 352·17 to 369·12 DDD/100 admissions). However, they observed a changed profile of antibiotic consumption: an increase in the use of ampicillin and gentamicin, and a decrease in the use of beta-lactamase inhibitors, macrolides and meropenem.

Two remaining studies used unique measures of units to express their results. A randomized control trial by Tagare et al. was looking at incidence of sepsis as the primary outcome, and did not report results in units that describe amounts of used antibiotics.^41^ They found no significant differences in sepsis incidence or mortality between the control (71 infants, sepsis incidence 25·4 %, mortality 2·8 %) and intervention group (69 infants, sepsis incidence 31·9 %, mortality 2·9 %), also in the VLBW infants subgroup (sepsis incidence 42·3 % and 59·3 %, mortality 3·8 % and 7·4 % for VLBW infants in control and intervention group, respectively). Ting et al. investigated how the proportion of infants with negative blood cultures receiving unnecessary prolonged antibiotic therapy (> three days) changed after the implementation of ASP.^42^ They compared a pre-ASP and a post-ASP cohort and found a reduction in inappropriate courses of therapy with meropenem, vancomycin, cefotaxim and linezolid.

### Discussion

Several approaches may reduce unnecessary antibiotic exposure for premature infants. In this systematic review we identified 11 articles using different approaches grouped under three action areas. We found reduction in use of antibiotics in studies focusing directly on reducing initiation or on shortening the duration of antibiotic therapy. Studies focusing solely on general intentions, without specific individual-dependent measures, did not demonstrate reduced antibiotic consumption. They were however able to achieve a reduction of the use of resistance driving broad prescribed antibiotics. We cannot statistically compare the studies but propose that specific action towards individual neonatal patients are more efficient than general improvements and revisions of antibiotic prescribing practises in reducing unnecessary antibiotic exposure in premature infants.

The studies in this review have different starting points regarding populations, means and in particular guidelines and practise for prescribing antibiotics. When the baseline is “Five days of antibiotics to all premature babies”, small efforts are needed for significant improvement. In departments with the most severely ill and fragile neonates and/or where several measures to restrict unnecessary antibiotics have already been implemented, it is harder to see positive development. Nzegwu et al., for instance, performed their ASP in a level IV NICU with an already low baseline antibiotic consumption.^40^

There is a general lack of information on ASPs in premature infants, especially < 34 weeks. Only three of the studies selected in our review focus exclusively on this populations. Most of the selected studies included less than 50 % premature infants, and for results different stages of prematurity were seldom reported separately (details in Table 1). The exceptions are Tagare et al.^41^, including only infants born at ≤ 36 weeks GA, and showing promising results also in VLBW subgroup, Tolia et al.^43^ including only VLBW infants (average GA 28 weeks) showing significant reduction in antibiotic consumption after 48 hours automatic stop implementation and Ting et al.^42^ including infants with GA ranging from 25 to 35 weeks and showing a reduction of inappropriate prescriptions after ASP implementation. However, a sub-analysis of VLBW infants separately did not reflect these findings.^42^ In all other studies, no separate results for different stages of prematurity or for VLBW infants were reported. It is thus not clear if their changes of antibiotic therapy reflect mostly term, late-premature, or more immature infants.

American Academy of Pediatrics (AAP) recently published two clinical reports on EOS management, differentiating between infants born before or after 35 weeks GA, as approach towards premature infants needs to be different.^7,8,46^ A widely used online tool (http://kp.org/eoscalc^45^) to estimate the risk of EOS in infants born ≥ 34 weeks GA has contributed to a significant reduction in unnecessary use of antibiotics in term and late preterm infants, as shown in a recent meta-analysis.^47^ A similar prediction model for infants born at < 34 weeks GA is to our knowledge not available on the internet, but other published protocols could help on deciding when to withhold initiation of antibiotics. Kitano et al. created new guidelines for initiation of antibiotic therapy, based on clinical status of mother and infant, sepsis score of the infant, blood culture results and time-progression of symptoms, and achieved a significant reduction in percentage of patients started on antibiotic therapy.^36^ No sub-analysis of these subgroups was performed. Evaluation of “Clinical status” and “progression of symptoms” in premature infants are the more challenging parts of the tool.

When it comes to shortening duration of antibiotic therapy, the AAP states: “When blood cultures are sterile, antibiotic therapy should be discontinued by 36 - 48 hours of incubation, unless there is clear evidence of site-specific infection”.^46^ Several studies showed that an automatic stop of antibiotics after 36 - 48 hours efficiently decreased unnecessary antibiotic exposure in premature infant. Two studies^16,43^ used this as the only intervention and found significant reduction in total antibiotic doses on individual levels in two cohorts with different gestational ages, and one more study implemented automatic stop as one of its main actions.^37^ Another study implemented automatic stop as their 2^nd^ intervention, after thoroughly revising and troubleshooting their antibiotic prescriptions routines.^38^ It was this specific action that significantly reduced antibiotics consumption. All studies did however require clinical evaluation, some as supplement to the stop order, all for continuous evaluation of need to reconsider treatment. This is further emphasised by the relatively low reliability of blood culture results.^48^

The quality of the clinician’s evaluation is dependent not only on skills, but a number of other factors. Sufficient human and material resources are needed to enable frequent examination and meticulous monitoring of premature infants. Cooperation with obstetricians is essential for providing exact information of the circumstances of preterm birth, infectious status of the mother and thereby evaluation of risk factors. Proximity and communication with laboratories, their efficacy and development for fast identification of infectious agents and their resistance profile influence the possibility and the timeline of making decisions based on biochemistry and microbiology results.^49,50^ This may explain some of the variance in use of antibiotics across different NICU.

Infection prevention and control (IPC) aims to protect patients and health workers from being harmed by avoidable infections.^51^ Positive outcomes of successful IPC programs result in lower incidence of healthcare related resistant infections and thus lower antibiotic prescription rates.^51^ IPC actions range from hand hygiene, visitor limitations, sterile equipment, and vaccination of health care workers to interventions related to infrastructure, number of health care workers and special isolation actions. ASPs implemented alongside IPC are more successful as when implemented alone.^52^ Thus ASPs need adaptation to local resources, infrastructure, routines and epidemiological challenges. A Dutch study illustrates the importance of IPC for successful ASP implementation. Despite the introduction of quality improvement programmes and antibiotic stewardship two Dutch Level III NICUs observed an increase in Gram-negative LOS in ELBW infants.^53^ Even though this could be partly explained by the increased proportion of more premature infants, another observation was remarkably low adherence to hand hygiene practises.

Established clinical practise often needs revisions based on data revealing causal relationship between exposures to given treatment and serious harm. One example of this is the use of supplemental oxygen in premature infants. It is definitely a vital supportive action, but discoveries of negative effects such as increased retinopathy of prematurity and bronchopulmonary dysplasia urged a change in previous practise and targeting lower SpO2 levels in VLBW infants.^54^ Recent research has revealed adverse effect of antibiotic treatment on health of premature infants.^21,22^ Additionally, antibiotic resistance is a fast increasing global challenge. Antibiotic therapy in early life increases carriage of antibiotic resistance genes and could contribute to increased antibiotic resistance in the population.^55^ This is however not the main argument to restrict the use of antibiotics in neonates. Public health policies addressing the global resistance problem are important, but not the main argument to restrict the use of antibiotics in this group. Individual adverse outcomes associated with antibiotic exposure of infants should be emphasized when deciding on initiating and discontinuing antibiotic treatment. A 25-years long prospective study in Australia showed that to stop empiric antibiotic treatment after 2-3 days in neonates (in the absence of positive blood cultures), subsequently reducing antibiotic consumption, did not prevent emergence of cephalosporin-resistant Gram-negative bacteria.^56^ Thus, attention must be given to negative consequences of antibiotic exposure on the individual level.

As it is more important to focus on individual consequences of antibiotic use and not the total antibiotic consumption within the department, reporting the number of days of spared antibiotic treatment per infant rather than non-individualized measure units such as DDD would improve the information in reported ASP results.

The clinician needs to balance the fear of not providing necessary antibiotics to effectively treat an infection with the risk of short and long-term negative effects. Local customized studies similar to the one from Tagare et al.^41^ are important to reduce the fear of the clinicians of overlooking need of antibiotics resulting in disastrous effects.

We further compared the decrease in antibiotic consumption that the studies were able to achieve, and the action areas they included in their ASP. Kitano et al. combined actions from all three different areas and achieved highest decrease in DOT/1000 patient-days.^36^ Studies combining two of the action areas presented lower decrease (28 – 30 %)^37,38,43^, while the articles focusing on one action area alone had the lowest, 25 %^16^ and 15 %^40^ decrease. On note, the study with the least decrease in antibiotic consumption had already implemented several actions from the other areas, had sicker patients and an already low antibiotic consumption.

Even though health behaviour interventions of ASP largely target clinical personnel, it is imperative that the unit, department and hospital leadership acknowledge the clinical challenges, encourage transparency and nonpunitive culture even when adverse outcome occurs, and endorse these programs.^57,58^ The study by Nzegwu et al. also demonstrates the fruits of joint efforts between health authorities and clinicians.^40^ Patel et al. modified aspects of the Get Smart for Healthcare campaign.^59^ They found numerous barriers when following the model of actionable feedback and provided a framework to design and implement an ASP mainly focusing on audit and feedback intervention for clinicians in a NICU. Nzewgu et al. later used these findings in their study.^40^

## Conclusion

In the reviewed studies, development and implementation of multivariable risk assessments and clinical tools for decisions on initiation of antibiotic treatment of suspected or potential sepsis, and using an automatic stop in antibiotic prescriptions appeared to be the most successful actions in reducing unnecessary antibiotic exposure in premature infants. A thorough evaluation of the current state at the NICU also helps identify weak points of antibiotic prescribing practises and allow for a custom tailored ASP.^38^ These actions are more demanding to implement in clinical practise than use of online calculator only, but suitable for infants with GA < 34 weeks. In reviewed studies, general actions for limiting antibiotic use on the hospital level only, were not successful in reducing unnecessary antibiotic exposure in premature infants, but could improve the profile of used antibiotics. This may lead to the presumption that a broad and multifactorial approach is preferable in ASP.

Due to the risk of adverse short and long-term effects of antibiotics on the developing infant it is imperative that nonessential exposure is avoided.^45,60,61^ ASPs are beneficial in this exercise. It is necessary and possible to closely monitor AB exposure of premature infants locally to early reveal changes with potential negative impacts on mortality and morbidity, and in the long run, to demonstrate the positive effects of preserving the premature microbiome.

## Limitations

There are limited studies regarding ASP for premature intensive care units. Many of the current papers reviewed here relate to neonatal intensive care units and include only a selected group of premature infants. Reviewed studies varied in gestational age of included infants, settings and outcome measures. No reports of adverse outcomes are documented.

## Data Availability

Not applicable. As this is a systematic literature review, no original data was created.

## Author contribution statement

FCP and URD conceived the presented idea. All authors developed the protocol and PR, KH, URD, and ODS performed the literature search. PR took the lead in writing the manuscript. All authors provided critical feedback and helped share the research, analysis and manuscript.

## Acknowledgments

We thank Marie Susanna Isachsen from the University of Oslo library for helping with the protocol for the literature search.

## Appendix

### Appendix 1:Literature search

Search date: 9 July 2019

We conducted separate searches in the following electronic databases to identify relevant studies: PubMed (1107 hits), Cochrane Database of Systematic Reviews (0 hits), Cochrane Central Register of Controlled Trials (95 hits), UpToDate (0 hits), National Institute for Health and Care Excellence (NICE) (0 hits), McMaster Plus (0 hits). Additional eight articles were obtained from reference lists.

Total number of hits: 1210

Number of hits after removal of duplicates: 1210

A second search was preformed 05.12.2019 and revealed no additional studies.

The protocol of this systematic review was not registered. Data extraction began before 1st of October 2019 and the protocol was therefore not eligible for submission in Prospero.

#### PubMed

The search strings below were combined as follows:

(1 OR 2) AND 3 and limited to English and Scandinavian languages

String 1

“Antimicrobial Stewardship”[Mesh] OR “Drug Resistance, Microbial/prevention and control”[Mesh] OR stewardship[title] OR ((antibiotic*[title] OR antibacterial[title] OR anti-bacterial[title] OR antimicrob*[title] OR antiinfective[title] OR anti-infective[title]) AND resistan*[title])

String 2

(“Drug Utilization Review”[Mesh] OR “Practice Patterns, Physicians’”[Mesh] OR “Drug Prescriptions”[Mesh:NoExp] OR (prescription*[title] OR prescrib*[title]) AND practice*[title])) AND (antibiotic*[title] OR antibacterial[title] OR anti-bacterial[title] OR antimicrob*[title] OR antiinfective[title] OR anti-infective[title] OR “Anti-Bacterial Agents”[Mesh] OR “Anti-Infective Agents”[Majr:NoExp] OR “Drug Resistance, Microbial”[Mesh])

String 3

“Infant, Newborn”[Mesh] OR “Intensive Care, Neonatal”[Mesh] OR “Intensive Care Units, Neonatal”[Mesh] OR “Neonatology”[Mesh] OR “Perinatology”[Mesh] OR “Infant, Newborn, Diseases”[Mesh] OR “Perinatal Care”[Mesh] OR “Obstetric Labor, Premature”[Mesh] OR newborn*[ti] OR neonat*[ti] OR perinat*[ti] OR prematur*[ti] OR preterm*[ti] OR nicu[ti] OR postnat*[ti] OR post-nat*[ti] OR (low[ti] AND (birthweight*[ti] OR weight*[ti]))

Comments:

[Mesh] = Medical Subject Headings

[Majr] = MeSH Major Topic.

[Majr:NoExp] = does not include

MeSH terms found below this term in the

MeSH hierarchy

#### Cochrane database of systematic reviews

Searched this database via PubMed. Used the PubMed search strategy (see below) and added:

AND “The Cochrane database of systematic reviews”[Jour]

#### Cochrane Central Register of Controlled Trials

Searced via “EBM Reviews - Cochrane Clinical Answers”, part of Ovid’s Evidence Based Medicine Reviews collection

1. ((premature* or preterm*) and (stewardship* or antibiotic* or antibacterial or anti-bacterial or antimicrob* or antiinfective or anti-infective)).ti.
2. limit 1 to (Danish or English or Norwegian or Swedish)
3. use cctr

#### UpToDate

Browsed UpToDate Topics:

Contents > Pediatrics > Neonatology

+ Searched separately:

- Stewardship
- Antibiotics AND premature

#### National Institute for Health and Care Excellence (NICE)

Browsed guidelines:

- Population groups > Infants and neonates
- Conditions and diseases > Infections > Sepsis

+ Searched separately:

- Stewardship
- Antibiotics AND premature

#### McMaster PLUS

We searched separately:

- “stewardship AND premature”
- «interventions AND antibiotics AND premature»
- “prescription practices AND premature”

### Appendix 2: Full text articles excluded

1. Alturk et al. (2018)^62^ looked at the factors responsible for prolonged antibiotic therapy in premature infants born < 29 weeks GA. Even though identifying factors that prolong treatment is important, they did not further explore if influencing this same factors in real settings would have any beneficial effect.
2. Ariffin et al. (2018)^63^ accessed the influence of a ward tailored (NICU) antibiotic policy by comparing causative agents of nosocomial bloodstream infections with those found in an adult intensive care unit. We excluded this study as it focused on microorganism found in NICU, and not on the antibiotic consumption in infants.
3. Bertini et al. (2013)^64^ evaluated an indirect action to reduce consumption of antibiotics by using special coated catheters for prevention of catheter-related bloodstream infections (CRBSI). Even though they observed a significant reduction in CRBSI (p = 0·005), there was no difference in consumption of antibiotic prophylaxis. They did also not report how lower rates of infections influenced antibiotic consumption at the NICU during the infants stay. We decided to exclude this study on the basis that their primary outcomes were not directed towards lowering consumption of antibiotics, but rather preventing infections.
4. De Man et al. (2000)^65^ implemented an antibiotic policy but reported its effects as emergence of resistant bacteria. They showed that policies regarding empiric antibiotic therapy influence the control of antimicrobial resistance, but reported no data on how the policy influenced antibiotic consumption for infants, and were thus excluded.
5. Di Pentima et al. (2010)^66^ described the impact of the implementation of an ASP on prescription errors for hospitalized children. They did not report the effect of ASP on use of antibiotics in infants, and were thus excluded.
6. Garner et al. (2015)^67^ evaluated the effectiveness of an interactive computerized order set to prevent prescription errors in neonatal late-onset sepsis, but did not evaluate the influence of this program on shortened or prolonged antibiotic therapy, length of stay or mortality rates.
7. Ho et al. (2018)^68^ only observed the adherence of ASP according to CDC recommendations, but did not report data on antibiotic consumption before/after implementation of this program.
8. Kuzniewicz et al. (2017)^45^ created a predictive model of neonatal EOS risk, including a study cohort of 204,485 infants born at GA ≥ 35 weeks. Their work has been an important milestone in reducing unnecessary exposure in premature infants. However, we decided not to include this paper in our review, as they did not report the numbers of premature infants (only the proportion of infants with GA < 38 weeks), and their average GA was > 39 weeks.
9. Malcolmson et al. (2017)^69^ investigated the combined impact of matrix-assisted laser desorption/ionization time-of-flight (MALDI-TOF) technology and an ASP in paediatric patients with blood stream infections (BSIs). We excluded their study as they focused on reducing the time to optimal antibiotic treatment, and did not report changes in initiation or duration of antibiotic treatment in infants.
10. Money et al. (2017)^70^ performed a hypothetical retrospective study to evaluate if the use of EOS calculator (developed by Kaiser Permanente) would safely reduce antibiotic use in well-appearing term infants born to mothers with chorioamnionitis. This hypothetical study showed that management according to the EOS calculator would reduce antibiotic use in infants (p = 0·0001) and average length of therapy (p = 0·0001). The study was excluded because the cohort was composed of term infants.
11. O’Leary et al. (2019)^71^ described a surveillance strategy to monitor antibiotic use and improve antibiotic stewardship in neonates. Their study was excluded due to lack of data on antibiotic use before and after implementation of this strategy.
12. Patel et al. (2009)^72^ only observed the adherence of ASP according to CDC recommendations, but did not report data on antibiotic consumption before/after implementation of this program.
13. Steinmann et al. (2018)^58^ assessed impact of empowering leadership style on ASP in a NICU/PICU over 3 years. They reported a significant decline in antibiotic days per 1,000 patient days. We excluded this study since it was targeted towards all paediatric patients and not specific to neonatal patients, and did not analyse data for neonatal patients separately.
14. Stocker et al. (2012)^73^ evaluated a three months surveillance strategy for antibiotic consumption according to the CDC 12-step capaign in a pediatric intensive care unit. They reported increassed percantage of appripriate empiric therapy courses (p < 0·001), increassed correct targeting of pathogen (p = 0·21) and reduced duration of therapy (p = 0·05). We excluded this study as it focused on all pediatric patients did not report any data seperatly for neonatal patients.
15. Toltzis et al. (2002)^74^ focused on the influence of antibiotic rotations in NICU (monthly rotation of gentamicin, piperacillin-tazobactam, and ceftazidime) on colonisation with resistant microorganism in the infants. The study was excluded as they did not report patent-level data on initiation and duration of antibiotic treatment.
16. Walker et al. (2017)^75^ showed results of ASP in neonatal surgical patients. This is a specific group of infants, mostly not premature, in specific situations (antibiotic prophylaxis at surgical procedures). We excluded the study as such ASP cannot be applied for sepsis in premature infants.

## References

1. Aminov RI. A brief history of the antibiotic era: lessons learned and challenges for the future. Front Microbiol. 2010;1:134.

2. Clatworthy AE, Pierson E, Hung DT. Targeting virulence: a new paradigm for antimicrobial therapy. Nat Chem Biol. 2007;3(9):541–8.

3. Armstrong GL, Conn LA, Pinner RW. Trends in infectious disease mortality in the United States during the 20th century. JAMA. 1999;281(1):61–6.

4. Superbugs threaten hospital patients. Center for Dissease Control and Prevention. Prevention.Atlanta, GA. March 2016 [Cited 10.12.2018]. Available from: https://www.cdc.gov/media/releases/2016/p0303-superbugs.html.

5. Lucia Hug DS, and Danzhen You. Levels & Trends in Child Mortality: UNICEF; 2017 [Cited 10.10.2019]. Available from: https://reliefweb.int/report/world/levels-and-trends-child-mortality-2017-report.

6. Klingenberg C, Kornelisse RF, Buonocore G, Maier RF, Stocker M. Culture-Negative Early-Onset Neonatal Sepsis - At the Crossroad Between Efficient Sepsis Care and Antimicrobial Stewardship. Frontiers in pediatrics. 2018;6:285. doi: 10.3389/fped.2018.00285.

7. Puopolo KM, Benitz WE, Zaoutis TE. Management of Neonates Born at >/=35 0/7 Weeks’ Gestation With Suspected or Proven Early-Onset Bacterial Sepsis. Pediatrics. 2018;142(6).

8. Puopolo KM, Benitz WE, Zaoutis TE. Management of Neonates Born at </=34 6/7 Weeks’ Gestation With Suspected or Proven Early-Onset Bacterial Sepsis. Pediatrics. 2018;142(6).

9. Schrag SJ, Farley MM, Petit S, Reingold A, Weston EJ, Pondo T, et al. Epidemiology of Invasive Early-Onset Neonatal Sepsis, 2005 to 2014. Pediatrics. 2016;138(6).

10. Weston EJ, Pondo T, Lewis MM, Martell-Cleary P, Morin C, Jewell B, et al. The burden of invasive early-onset neonatal sepsis in the United States, 2005-2008. Pediatr Infect Dis J. 2011;30(11):937–41.

11. Stoll BJ, Hansen NI, Sanchez PJ, Faix RG, Poindexter BB, Van Meurs KP, et al. Early onset neonatal sepsis: the burden of group B Streptococcal and E. coli disease continues. Pediatrics. 2011;127(5):817–26.

12. Stoll BJ, Hansen NI, Bell EF, Walsh MC, Carlo WA, Shankaran S, et al. Trends in Care Practices, Morbidity, and Mortality of Extremely Preterm Neonates, 1993-2012. JAMA. 2015;314(10):1039–51.

13. Puopolo KM, Draper D, Wi S, Newman TB, Zupancic J, Lieberman E, et al. Estimating the probability of neonatal early-onset infection on the basis of maternal risk factors. Pediatrics. 2011;128(5):e1155–63.

14. Dyar OJ, Huttner B, Schouten J, Pulcini C. What is antimicrobial stewardship? Clin Microbiol Infect. 2017;23(11):793–8.

15. MacDougall C, Polk RE. Antimicrobial stewardship programs in health care systems. Clin Microbiol Rev. 2005;18(4):638–56.

16. Astorga MC, Piscitello KJ, Menda N, Ebert AM, Ebert SC, Porte MA, et al. Antibiotic Stewardship in the Neonatal Intensive Care Unit: Effects of an Automatic 48-Hour Antibiotic Stop Order on Antibiotic Use. Journal of the Pediatric Infectious Diseases Society. 2019;25;8(4):310–316.

17. Hsieh EM, Hornik CP, Clark RH, Laughon MM, Benjamin DK, Jr., Smith PB, et al. Medication use in the neonatal intensive care unit. Am J Perinatol. 2014;31(9):811–21.

18. Cantey JB, Wozniak PS, Pruszynski JE, Sanchez PJ. Reducing unnecessary antibiotic use in the neonatal intensive care unit (SCOUT): a prospective interrupted time-series study. Lancet Infect Dis. 2016;16(10):1178–84.

19. Ting JY, Synnes A, Roberts A, Deshpandey A, Dow K, Yoon EW, et al. Association Between Antibiotic Use and Neonatal Mortality and Morbidities in Very Low-Birth-Weight Infants Without Culture-Proven Sepsis or Necrotizing Enterocolitis. JAMA pediatrics. 2016;170(12):1181–7.

20. Esaiassen E, Fjalstad JW, Juvet LK, van den Anker JN, Klingenberg C. Antibiotic exposure in neonates and early adverse outcomes: a systematic review and meta-analysis. J Antimicrob Chemother. 2017;72(7):1858–70.

21. Cotten CM, Taylor S, Stoll B, Goldberg RN, Hansen NI, Sanchez PJ, et al. Prolonged duration of initial empirical antibiotic treatment is associated with increased rates of necrotizing enterocolitis and death for extremely low birth weight infants. Pediatrics. 2009;123(1):58–66.

22. Alexander VN, Northrup V, Bizzarro MJ. Antibiotic exposure in the newborn intensive care unit and the risk of necrotizing enterocolitis. J Pediatr. 2011;159(3):392–7.

23. Groer MW, Luciano AA, Dishaw LJ, Ashmeade TL, Miller E, Gilbert JA. Development of the preterm infant gut microbiome: a research priority. Microbiome. 2014;2:38.

24. Trasande L, Blustein J, Liu M, Corwin E, Cox LM, Blaser MJ. Infant antibiotic exposures and early-life body mass. International journal of obesity (2005). 2013;37(1):16–23.

25. Marra F, Lynd L, Coombes M, Richardson K, Legal M, FitzGerald JM, et al. Does Antibiotic Exposure During Infancy Lead to Development of Asthma?: A Systematic Review and Metaanalysis. Chest. 2006;129(3):610–8.

26. Vangay P, Johnson AJ, Ward TL, Al-Ghalith GA, Shields-Cutler RR, Hillmann BM, et al. US Immigration Westernizes the Human Gut Microbiome. Cell. 2018;175(4):962-72.e10.

27. McDonald B, McCoy KD. Maternal microbiota in pregnancy and early life. Science. 2019;365(6457):984.

28. Langdon A, Crook N, Dantas G. The effects of antibiotics on the microbiome throughout development and alternative approaches for therapeutic modulation. Genome Med. 2016;8(1):39-.

29. Gasparrini AJ, Crofts TS, Gibson MK, Tarr PI, Warner BB, Dantas G. Antibiotic perturbation of the preterm infant gut microbiome and resistome. Gut Microbes. 2016;7(5):443–9.

30. Flannery DD, Ross RK, Mukhopadhyay S, Tribble AC, Puopolo KM, Gerber JS. Temporal Trends and Center Variation in Early Antibiotic Use Among Premature Infants. JAMA Network Open. 2018;1(1):e180164–e.

31. Norwegian Neonatal Network, Norwegian Institute of Public Health. [Cited 27.08.2019]. Available from: http://www.kvalitetsregistre.no/resultater/skade-og-inten-sivbehandling/norsk-nyfoedtmedisinsk-kvalitetsregister/.

32. Broadfoot M. A delicate balance. Science. 2018;360(6384):18–20.

33. Shamseer L, Moher D, Clarke M, Ghersi D, Liberati A, Petticrew M, et al. Preferred reporting items for systematic review and meta-analysis protocols (PRISMA-P) 2015: elaboration and explanation. BMJ : British Medical Journal. 2015;349:g7647.

34. Achten NB, Dorigo-Zetsma JW, van der Linden PD, van Brakel M, Plotz FB. Sepsis calculator implementation reduces empiric antibiotics for suspected early-onset sepsis. Eur J Pediatr. 2018;177(5):741–6.

35. Jinka DR, Gandra S, Alvarez-Uria G, Torre N, Tadepalli D, Nayakanti RR. Impact of Antibiotic Policy on Antibiotic Consumption in a Neonatal Intensive Care Unit in India. Indian Pediatr. 2017;54(9):739–41.

36. Kitano T, Takagi K, Arai I, Yasuhara H, Ebisu R, Ohgitani A, et al. A simple and feasible antimicrobial stewardship program in a neonatal intensive care unit of a Japanese community hospital. Journal of infection and chemotherapy : official journal of the Japan Society of Chemotherapy. 2019.

37. Lu C, Liu Q, Yuan H, Wang L. Implementation of the Smart Use of Antibiotics Program to Reduce Unnecessary Antibiotic Use in a Neonatal ICU: A Prospective Interrupted Time-Series Study in a Developing Country. Crit Care Med. 2019;47(1):e1–e7.

38. McCarthy KN, Hawke A, Dempsey EM. Antimicrobial stewardship in the neonatal unit reduces antibiotic exposure. Acta Paediatr. 2018;107(10):1716–21.

39. Nitsch-Osuch A, Kurpas D, Kuchar E, Zycinska K, Zielonka T, Wardyn K. Antibiotic consumption pattern in the neonatal special care unit before and after implementation of the hospital’s antibiotic policy. Adv Exp Med Biol. 2015;835:45–51.

40. Nzegwu NI, Rychalsky MR, Nallu LA, Song X, Deng Y, Natusch AM, et al. Implementation of an Antimicrobial Stewardship Program in a Neonatal Intensive Care Unit. Infect Control Hosp Epidemiol. 2017;38(10):1137–43.

41. Tagare A, Kadam S, Vaidya U, Pandit A. Routine antibiotic use in preterm neonates: a randomised controlled trial. J Hosp Infect. 2010;74(4):332–6.

42. Ting JY, Paquette V, Ng K, Lisonkova S, Hait V, Shivanada S, et al. Reduction of Inappropriate Antimicrobial Prescriptions in a Tertiary Neonatal Intensive Care Unit After Antimicrobial Stewardship Care Bundle Implementation. Pediatr Infect Dis J. 2019;38(1):54–9.

43. Tolia VN, Desai S, Qin H, Rayburn PD, Poon G, Murthy K, et al. Implementation of an Automatic Stop Order and Initial Antibiotic Exposure in Very Low Birth Weight Infants. Am J Perinatol. 2017;34(2):105–10.

44. Guillot J, Lebel D, Roy H, Ovetchkine P, Bussières J-F. Usefulness of defined daily dose and days of therapy in pediatrics and obstetrics-gynecology: a comparative analysis of antifungal drugs (2000-2001, 2005-2006, and 2010-2011). J Pediatr Pharmacol Ther. 2014;19(3):196–201.

45. Kuzniewicz MW, Puopolo KM, Fischer A, Walsh EM, Li S, Newman TB, et al. A Quantitative, Risk-Based Approach to the Management of Neonatal Early-Onset Sepsis. JAMA pediatrics. 2017;171(4):365–71.

46. Puopolo KM. New sepsis guidance adresses epidemiology, microbiology, recommended empiric treatment AAP News2018. [Cited 13.01.2020]. Available from:https://www.aappublications.org/news/2018/11/19/sepsis111918?utm_source=TrendMD&utm_medium=TrendMD&utm_campaign=AAPNews_TrendMD_0.

47. Achten NB, Klingenberg C, Benitz WE, et al. Association of Use of the Neonatal Early-Onset Sepsis Calculator With Reduction in Antibiotic Therapy and Safety: A Systematic Review and Meta-analysis. JAMA Pediatr. 2019;173(11):1032– 1040. doi:10.1001/jamapediatrics.2019.2825.

48. Zea-Vera A, Ochoa TJ. Challenges in the diagnosis and management of neonatal sepsis. J Trop Pediatr. 2015;61(1):1–13.

49. Romaniszyn D, Rozanska A, Wojkowska-Mach J, Chmielarczyk A, Pobiega M, Adamski P, et al. Epidemiology, antibiotic consumption and molecular characterisation of Staphylococcus aureus infections--data from the Polish Neonatology Surveillance Network, 2009-2012. BMC Infect Dis. 2015;15:169.

50. Paul SP, Caplan EM, Morgan HA, Turner PC. Barriers to implementing the NICE guidelines for early-onset neonatal infection: cross-sectional survey of neonatal blood culture reporting by laboratories in the UK. J Hosp Infect. 2018;98(4):425–8.

51. WHO. Infection prevention and control [Cited 2019 31.05.2019]. Available from: https://www.who.int/gpsc/ipc/en/.

52. Baur D, Gladstone BP, Burkert F, Carrara E, Foschi F, Dobele S, et al. Effect of antibiotic stewardship on the incidence of infection and colonisation with antibiotic-resistant bacteria and Clostridium difficile infection: a systematic review and meta-analysis. Lancet Infect Dis. 2017;17(9):990–1001.

53. Ran NC, van den Hoogen A, Hemels MAC. Gram-negative Late-onset Sepsis in Extremely Low Birth Weight Infants Is Emerging in The Netherlands Despite Quality Improvement Programs and Antibiotic Stewardship! The Pediatric Infectious Disease Journal. 2019;38(9):952–7.

54. Saugstad OD, Aune D. In search of the optimal oxygen saturation for extremely low birth weight infants: a systematic review and meta-analysis. Neonatology. 2011;100(1):1–8.

55. Gasparrini AJ, Wang B, Sun X, Kennedy EA, Hernandez-Leyva A, Ndao IM, et al. Persistent metagenomic signatures of early-life hospitalization and antibiotic treatment in the infant gut microbiota and resistome. Nature Microbiology. 2019;4(12):2285–97.

56. Carr D, Barnes EH, Gordon A, Isaacs D. Effect of antibiotic use on antimicrobial antibiotic resistance and late-onset neonatal infections over 25 years in an Australian tertiary neonatal unit. Arch Dis Child Fetal Neonatal Ed. 2017;102(3):F244–f50.

57. Lawrence KL, Kollef MH. Antimicrobial stewardship in the intensive care unit: advances and obstacles. Am J Respir Crit Care Med. 2009;179(6):434–8.

58. Steinmann KE, Lehnick D, Buettcher M, Schwendener-Scholl K, Daetwyler K, Fontana M, et al. Impact of Empowering Leadership on Antimicrobial Stewardship: A Single Center Study in a Neonatal and Pediatric Intensive Care Unit and a Literature Review. Frontiers in pediatrics. 2018;6:294.

59. Patel SJ, Saiman L, Duchon JM, Evans D, Ferng YH, Larson E. Development of an antimicrobial stewardship intervention using a model of actionable feedback. Interdiscip Perspect Infect Dis. 2012;2012:150367.

60. Puopolo KM, Mukhopadhyay S, Hansen NI, Cotten CM, Stoll BJ, Sanchez PJ, et al. Identification of Extremely Premature Infants at Low Risk for Early-Onset Sepsis. Pediatrics. 2017;140(5).

61. Mukhopadhyay S, Puopolo KM. Clinical and Microbiologic Characteristics of Early-onset Sepsis Among Very Low Birth Weight Infants: Opportunities for Antibiotic Stewardship. The Pediatric infectious disease journal. 2017;36(5):477–81.

62. Alturk MR, Baier RJ. Patient and prescriber factors and the prolongation of antibiotics after birth in infants less than 29 weeks. J Matern Fetal Neonatal Med. 2018;31(13):1720–6.

63. Ariffin N, Hasan H, Ramli N, Ibrahim NR, Taib F, Rahman AA, et al. Comparison of antimicrobial resistance in neonatal and adult intensive care units in a tertiary teaching hospital. Am J Infect Control. 2012;40(6):572–5.

64. Bertini G, Elia S, Ceciarini F, Dani C. Reduction of catheter-related bloodstream infections in preterm infants by the use of catheters with the AgION antimicrobial system. Early Hum Dev. 2013;89(1):21–5.

65. de Man P, Verhoeven BA, Verbrugh HA, Vos MC, van den Anker JN. An antibiotic policy to prevent emergence of resistant bacilli. Lancet. 2000;355(9208):973–8.

66. Di Pentima MC, Chan S. Impact of antimicrobial stewardship program on vancomycin use in a pediatric teaching hospital. Pediatr Infect Dis J. 2010;29(8):707–11.

67. Garner SS, Cox TH, Hill EG, Irving MG, Bissinger RL, Annibale DJ. Prospective, controlled study of an intervention to reduce errors in neonatal antibiotic orders. J Perinatol. 2015;35(8):631–5.

68. Ho T, Buus-Frank ME, Edwards EM, Morrow KA, Ferrelli K, Srinivasan A, et al. Adherence of Newborn-Specific Antibiotic Stewardship Programs to CDC Recommendations. Pediatrics. 2018;142(6).

69. Malcolmson C, Ng K, Hughes S, Kissoon N, Schina J, Tilley PA, et al. Impact of Matrix-Assisted Laser Desorption and Ionization Time-of-Flight and Antimicrobial Stewardship Intervention on Treatment of Bloodstream Infections in Hospitalized Children. Journal of the Pediatric Infectious Diseases Society. 2017;6(2):178–86.

70. Money N, Newman J, Demissie S, Roth P, Blau J. Anti-microbial stewardship: antibiotic use in well-appearing term neonates born to mothers with chorioamnionitis. J Perinatol. 2017;37(12):1304–9.

71. O’Leary EN, van Santen KL, Edwards EM, Braun D, Buus-Frank ME, Edwards JR, et al. Using NHSN’s Antimicrobial Use Option to Monitor and Improve Antibiotic Stewardship in Neonates. Hospital pediatrics. 2019;9(5):340–7.

72. Patel SJ, Oshodi A, Prasad P, Delamora P, Larson E, Zaoutis T, et al. Antibiotic use in neonatal intensive care units and adherence with Centers for Disease Control and Prevention 12 Step Campaign to Prevent Antimicrobial Resistance. Pediatr Infect Dis J. 2009;28(12):1047–51.

73. Stocker M, Ferrao E, Banya W, Cheong J, Macrae D, Furck A. Antibiotic surveillance on a paediatric intensive care unit: easy attainable strategy at low costs and resources. BMC Pediatr. 2012;12:196.

74. Toltzis P, Dul MJ, Hoyen C, Salvator A, Walsh M, Zetts L, et al. The effect of antibiotic rotation on colonization with antibiotic-resistant bacilli in a neonatal intensive care unit. Pediatrics. 2002;110(4):707–11.

75. Walker S, Datta A, Massoumi RL, Gross ER, Uhing M, Arca MJ. Antibiotic stewardship in the newborn surgical patient: A quality improvement project in the neonatal intensive care unit. Surgery. 2017;162(6):1295–303.

